# Predictors of non-participation in azithromycin mass drug administration to reduce mortality among children 1-11 months old in Niger: a coverage evaluation survey

**DOI:** 10.64898/2026.02.04.26345612

**Authors:** Carolyn Brandt, Ahmed M. Arzika, Abdou Amza, Ramatou Maliki, Alio Mankara, Nasser Gallo, Abdoul Naser Harouna, Diallo Beidi, Elodie Lebas, Brittany Peterson, Benjamin F. Arnold, Thomas M. Lietman, Kieran S. O’Brien, the AVENIR Study Group

## Abstract

The World Health Organization recommends biannual azithromycin mass drug administration (MDA) to infants aged 1-11 months to reduce mortality, following promising results from trials in West Africa. High coverage seen in well-resourced trials may decline as the intervention transitions to a real-world program. As a result, the most vulnerable children facing the highest risk of mortality may be missed. We aimed to identify predictors of non-participation in an azithromycin MDA program to inform programmatic delivery strategies to improve coverage. We conducted a coverage evaluation survey after azithromycin MDA to children aged 1-11 months in Niger’s Tahoua region. Data collection teams visited households to assess caregiver-reported participation, reasons for participation and non-participation, and adverse events. Mixed effects logistic regression models were used to analyze community-, household-, and child-level predictors associated with non-participation in azithromycin MDA. Among 40 communities with 811 unique households and 871 children ages 2-12 months old included in analyses, 76% of eligible children received treatment based on caregiver report compared to 96% community health worker-reported coverage. The most frequently stated reasons for non-receipt of treatment were absence (34%), nobody coming to the house (31%), and not receiving enough information (17.2%). In an adjusted model, older children experienced higher odds of receiving treatment (aOR 1.22, 95% CI 1.15 - 1.30, *P* ≤ 0.0001), as did children living in more densely populated areas (aOR 1.15, 95% CI 1.04 - 1.28, *P* = 0.01). Adverse events were reported among 6.8% of children who received treatment, with fever being the most reported symptom. Strengthening community sensitization and preparation activities before MDA is essential to address common reasons for non-participation. Future research to understand why younger children and those living in sparsely populated communities were less likely to be included may help target specific interventions in these populations.

## Introduction

The World Health Organization recommends biannual azithromycin mass drug administration (MDA) to infants aged 1-11 months to reduce mortality in high mortality settings in sub-Saharan Africa, following the results of the MORDOR trial.[1]The mechanism of effect is thought to include both direct effects on children with respiratory infections, diarrhea, and malaria,[2] as well as indirect effects through reduction in community transmission of infectious diseases that can be treated with azithromycin.[3] Although specific coverage targets have not yet been set by the WHO for this intervention, high coverage is likely needed to achieve mortality reductions given the indirect effect mechanism. Coverage targets for MDA programs for neglected tropical diseases such as trachoma or onchocerciasis are 80% or greater.[4]

Evidence suggests that individuals missed in MDA programs are often the most vulnerable and may face a higher risk of mortality. In the MORDOR trial, data suggested that mortality rates among children missed by the MDA team were 1.5-2 times higher than those included in the intervention.[5] Prior studies exploring characteristics of those missed by lymphatic filariasis and soil transmitted helminths MDA programs have found higher proportions of missed persons in remote areas,[6,7] as well as among those with a lack of information on the intervention.[8] However, most prior coverage studies assessed MDA programs targeting children and adults, or only adults. As azithromycin MDA for child survival only includes young children, there may be unique factors associated with non-participation in this setting.

The AVENIR (*Azithromycine pour la Vie des Enfants au Niger: Implementation et Recherche*) Programmatic Trial aimed to transition azithromycin MDA for child survival in Niger from a resource-intensive trial setting to a programmatic real-world setting. As this intervention transitions to a program, treatment coverage may decrease as programs typically have fewer resources for supervision and follow-up. Therefore, monitoring activities included a coverage evaluation survey after the second MDA. The primary objective of the survey was to identify predictors of non-participation in the program, with factors such as shorter household distance to the primary health center and receipt of information about the intervention hypothesized to increase treatment uptake (participation).

## Methods

### Study Design

The AVENIR Programmatic Trial, conducted at the same time as the associated primary AVENIR mortality trial, was a cluster-randomized trial in the Tahoua and Maradi regions in Niger, where 1500 primary health center (Centre de Santé Integré, CSI) catchment areas were randomized 2:1 to receive biannual oral azithromycin distribution (20 mg/kg) to children aged 1-11 months (World Health Organization (WHO) guideline at the time) or no azithromycin distribution with normal care services. Normal care by community health workers in this setting includes prevention, screening, treatment of diseases such as malaria and pneumonia, nutritional advice, referral to health center, vaccination, and administration of vitamin A. Every 6 months over 1 year, sensitization campaigns were conducted to inform communities of the timing of the MDA and azithromycin was delivered door-to-door by existing community health workers (CHWs). During the second distribution in August 2023, azithromycin MDA was delivered to children 1-11 months of age across 42 primary health centers in the Tahoua region, and within 4 weeks of the MDA, a separate data collection team conducted a cross-sectional coverage evaluation survey, which explored caregiver-reported participation in the program, reasons for participation and non-participation, and adverse events.

### Ethical approval

Institutional Review Boards (IRB) at the Niger Ministry of Health (*Comité Nationale Éthique pour la Recherche en Santé*, N 041/2020/CNERS) and the University of California, San Francisco (19-28287) provided ethical approval for the trial, which included this survey. Verbal consent was obtained from regional, district, health center, and community leaders for all study activities. At the household level, verbal consent was obtained from the head of household or the caregiver using an informed consent script prior to the initiation of the survey. The AVENIR trial was overseen by a Data Safety and Monitoring Committee and was registered on clinicaltrials.gov (NCT05288023).

### Setting, eligibility, and participants

Community eligibility for the trial included safe and accessible location in the Tahoua or Maradi regions and verbal consent from regional, district, health center, and community leaders. Individual eligibility for the trial included residence in a study community, age 1-11 months, and caregiver informed verbal consent. Selection for the survey involved simple, random sampling of CSIs in the Tahoua region that had received their second MDA as part of the trial. Overall, 7 CSIs were selected, and within each CSI, all villages and households were included in the survey. Households were visited door-to-door by a trained data collection team within 4 weeks of the MDA. After obtaining verbal consent from the head of household, the team collected socio-demographic information on the head(s) of household and caregivers(s), and the survey continued if there are children 1-11-months old living in the household. After data collection, children who were 0-1 months of age were excluded from analyses since they would not have been born at the time of the azithromycin distribution, due to the time between distribution and the coverage survey.

### Data collection tools

The survey was adapted from coverage evaluation survey guidance for preventive chemotherapy for neglected tropical diseases from the WHO.[4] Questions were modified and added to assess delivery-related scenarios and adverse events specific to this intervention (Supplementary Table 1). The survey was translated from English to French and administered electronically using the Commcare application between August 28^th^, 2023 and September 5^th^, 2023. (Dimagi, Cambridge, MA USA). Multilingual study team members reviewed English and French questionnaires to confirm translation accuracy, and each question was translated into relevant local languages, including Zarma, Hausa, and Peul. Data collectors were trained on the study procedures and the survey instrument prior to visiting the communities. Each worker was equipped with a smartphone with the Commcare mobile application, a verbal informed consent script, personal protective equipment, and a bottle of azithromycin for caregiver reference. To ensure that the whole community was surveyed, maps of data collection progress in each community were reviewed to identify households that were missed.

### Variables collected

The primary outcome of interest was caregiver-reported treatment coverage. For each child under 12 months of age caregivers were asked whether the child received azithromycin as part of the MDA from the prior month. Caregiver-reported coverage was calculated as the proportion of eligible children reported to have received azithromycin at the most recent MDA. Administrative coverage was calculated using treatment numbers reported by CHWs as the numerator and CSI population estimates as the denominator. Other variables of interest included receipt of information about the distribution before it took place, primary reason for participation and non-participation in MDA, when and how information about the intervention was received, and experience of adverse events.

Independent variables included community-, household-, and child-level predictors of participation in azithromycin MDA. Child-level predictors included age, sex, and timing of receipt of information on the distribution (less than a week before, more than a week before, never received, or do not know). Household-level predictors included distance to nearest primary health care center, head of household education level (none, literate, higher education), number of members in household, and number of children under 12 months in a household. Community-level predictors included the number of households within a community, community mean child age, community distance to nearest primary health care center, and population density. Population density was calculated using a nearest neighbor analysis approach. For each household in the dataset with GPS coordinates, distances and indices of the 11 nearest household neighbors (including the household itself) were calculated. Next, the mean distance among each household’s 10 nearest neighbors (excluding itself) were calculated, and then all mean estimates in a community were averaged to produce a community-level indicator. Each community value was assigned a percentile rank, with the largest values getting the smallest ranks, meaning that lower percentiles correspond to less densely populated and higher percentiles are more densely populated.

### Sample size and sampling

Target coverage levels have not been set by the WHO for this intervention, but 80% is used for similar trachoma azithromycin MDA programs.[4] To estimate a target coverage of 80%, we estimated that including 7 CSIs, 49 grappes (7 average grappes per CSI), and 1,274 children under age 12-months old (26 average per grappe) would enable a precision of ±2.4% at the regional level, and ±6.0% at the CSI level.

### Statistical methods

Continuous community-, household-, and child-level predictors and the outcomes of interest were assessed descriptively using median and interquartile range (IQR) for continuous variables and frequencies and percentages for categorical variables, overall and stratified by CSI. Unadjusted mixed effects logistic regression models were used to analyze predictors associated with non-participation in azithromycin MDA, with random effects at the community-level to account for clustering. Predictors found to be associated with participation based on having a *P-* value of <0.10 were included in an adjusted mixed effects logistic regression model. Receipt of information about the azithromycin distribution was determined to be on the causal pathway between other predictors and participation in azithromycin MDA and so was investigated as a secondary outcome using a similar approach. After reviewing results, one CSI was excluded from analyses due to unreliable coverage numbers (0% across numerous villages determined by the study team to be inaccurate). A sensitivity analysis including this CSI along with children zero to one month of age was conducted to determine the consistency of the results. Analyses were conducted using R software (version 4.4.3). Two-sided *P*-values < 0.05 were considered statistically significant in adjusted models.

## Results

A total of 42 CSIs were included in the MDA, and 7 were randomly selected for participation in the coverage evaluation survey. This included a total of 57 secure communities where 3,851 households were contacted (Fig 1). Among these, 3,848/3,851 households consented to participate in the survey. After excluding CSI Abalak, 40 communities with 811 unique households and 871 children ages 2-12 months old were analyzed.

**Fig 1.**
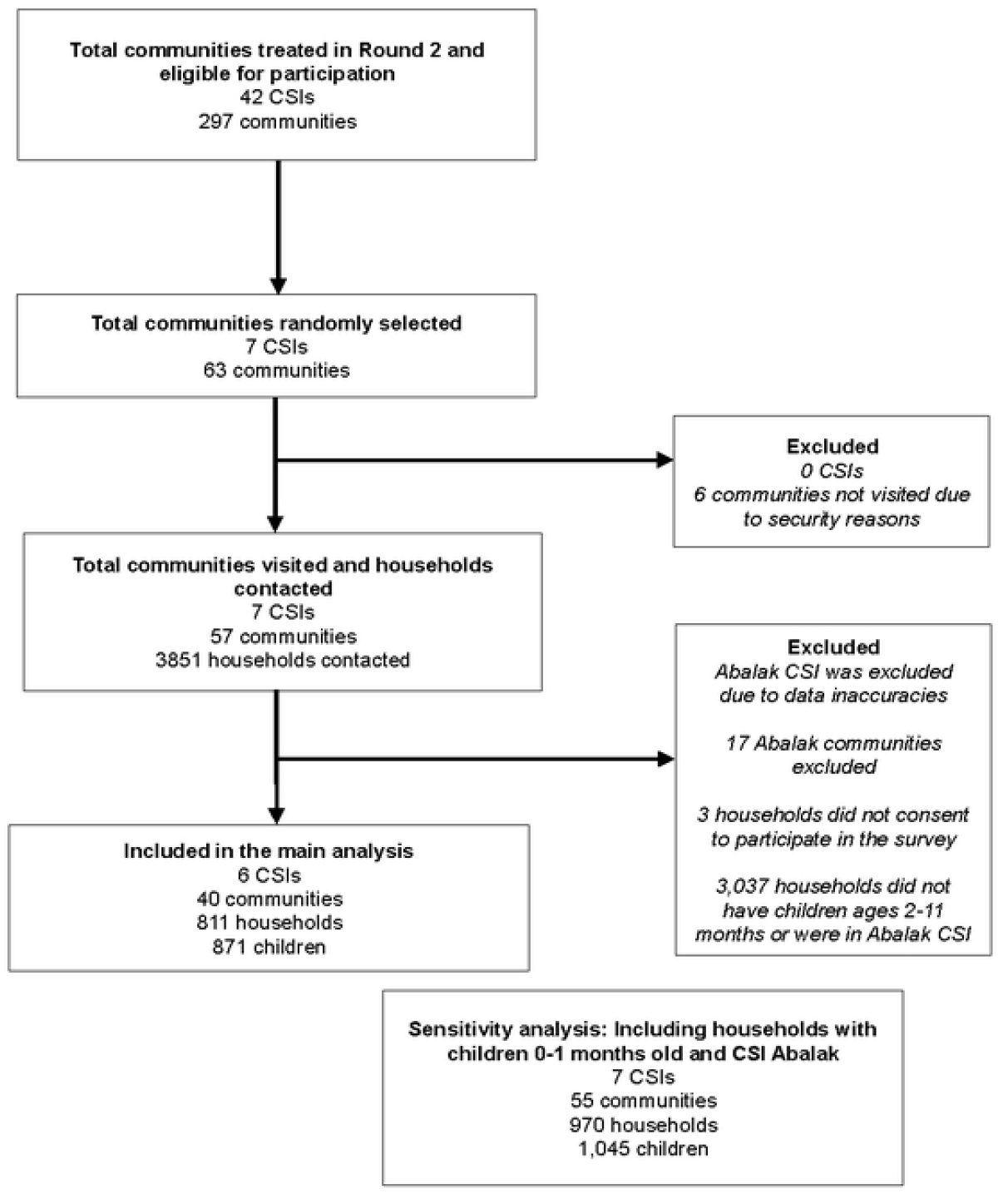
Participant Flow Diagram.

Table 1 presents the characteristics of survey respondents. Overall, 96.2% (838/871) of respondents with children eligible for treatment indicated that they were present for the azithromycin distribution, and 75.5% (658/871) stated that their eligible child received treatment (Table 1). Coverage varied by CSI (Fig 2, Supplementary Table 2), with a mean CSI-level caregiver-reported coverage of 71.5% (SD 10.8%, Supplementary Table 5). Administrative coverage among the surveyed CSIs was higher than caregiver-reported coverage both overall (Mean 96.0%, SD 4.2%) and for each CSI (Supplementary Table 5). Among those who did not receive treatment, the top three mutually exclusive reasons for non-receipt were being absent (34%, 69/203), nobody coming to the house (31%, 63/203), and not receiving enough information (17.2%, 35/203). Among children who did receive treatment, 6.8% (45/658) experienced any type of adverse event overall, with the most common being fever (48.9%, 22/45) diarrhea (31.1%, 14/45), and vomiting (17.8%, n=8/45) (Table 1).

**Table 1.**
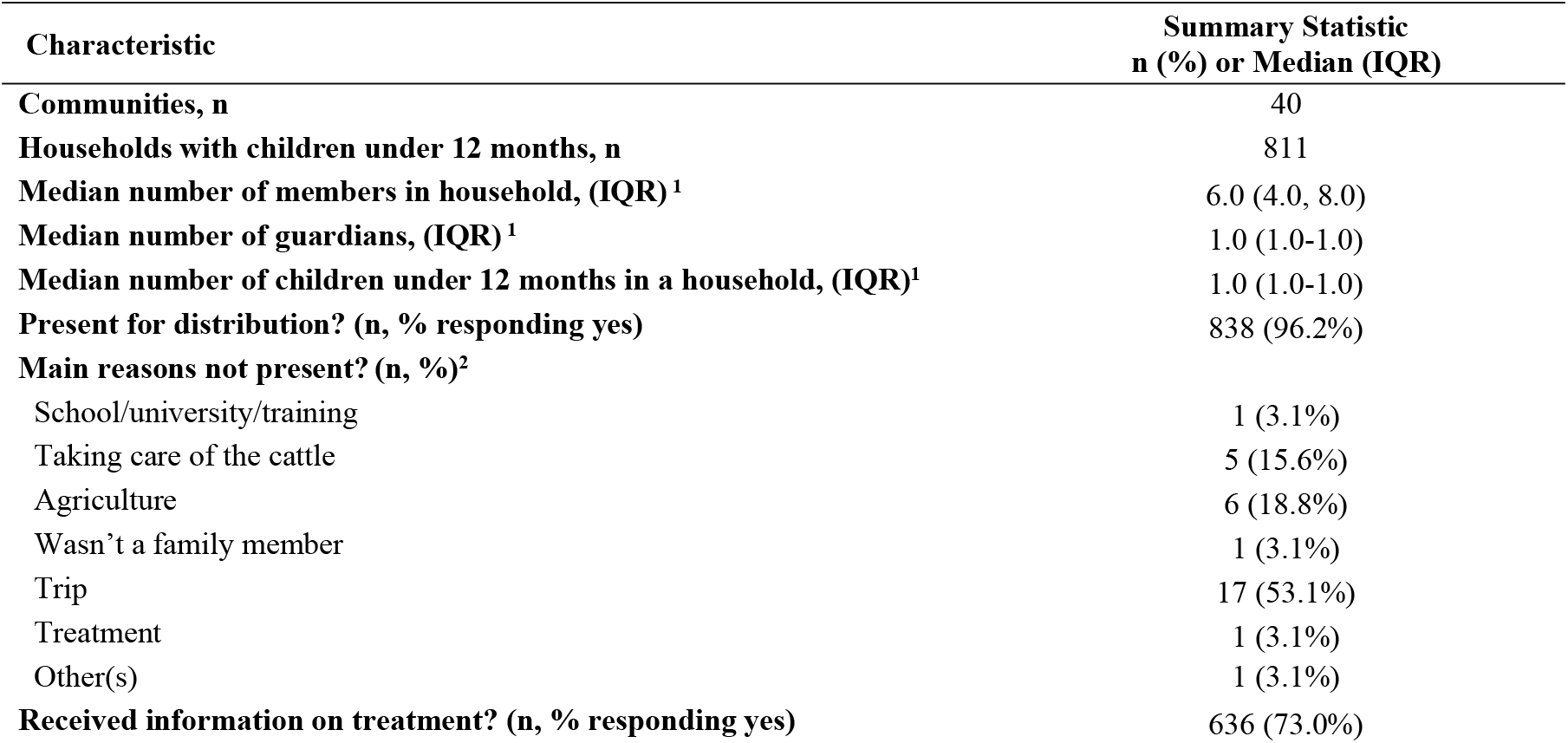
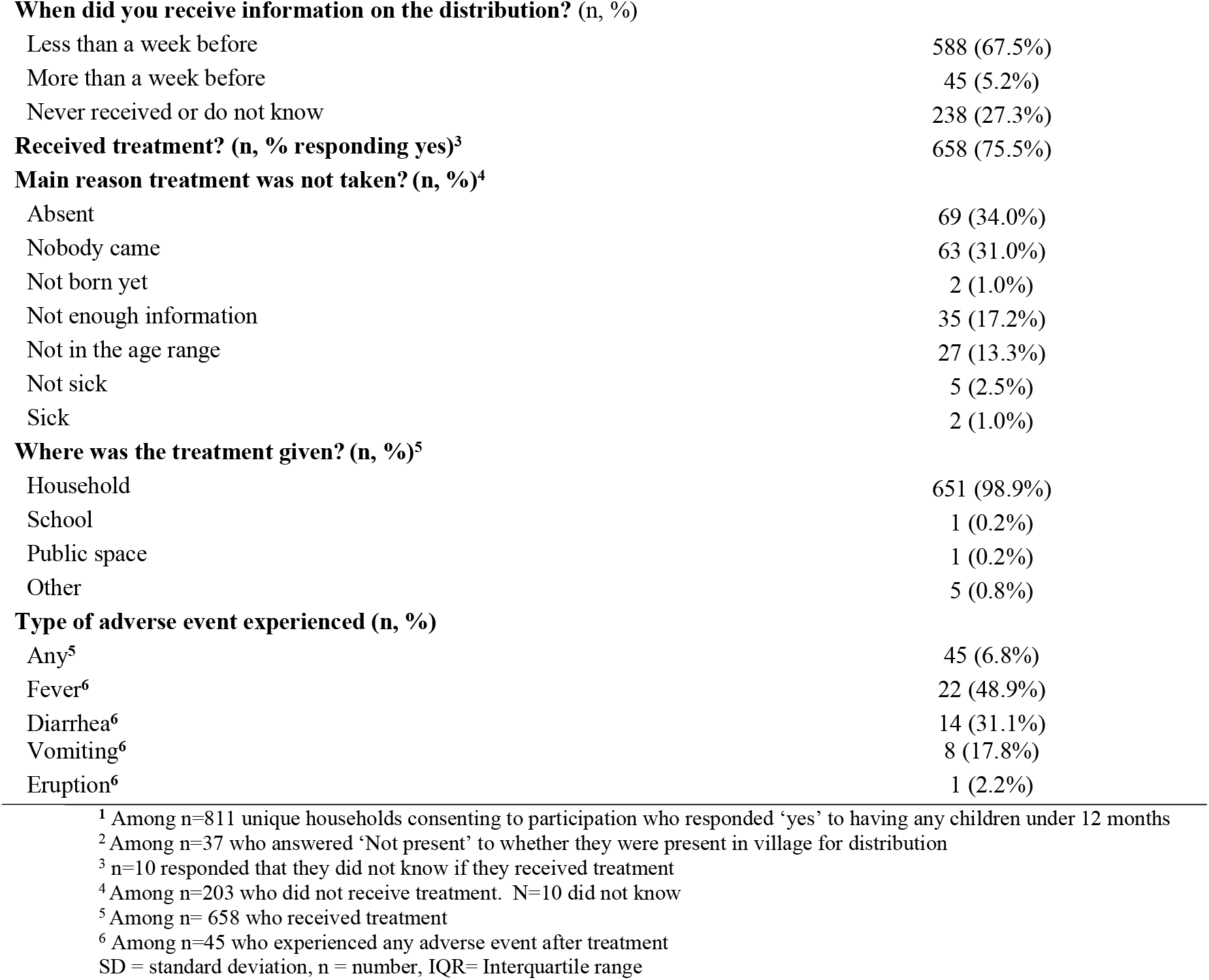
Descriptive characteristics of included survey responses among households with at least one child under 12 months old (n=871). CSI Abalak and children 0-1 months of age were excluded.

**Fig 2.**
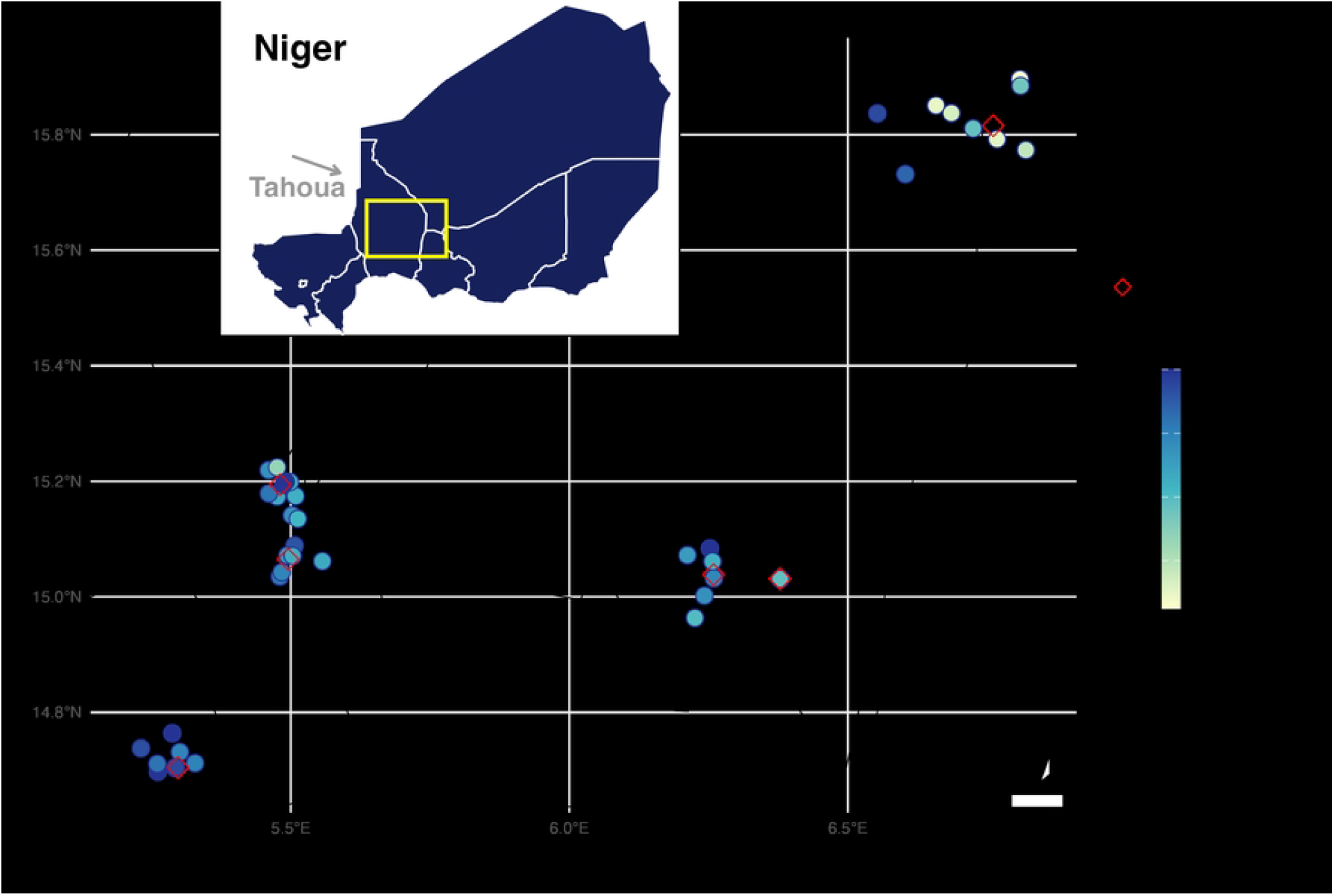
Community level treatment coverage, across 6 CSIs. Geographic distribution of six CSIs (red diamonds) and 40 communities (circles) in the Tahoua region of Niger. The community circle colors indicate the community-level treatment coverage percentage.

Age, education, and population density were found to be associated with caregiver-reported coverage (Table 2, Fig 3). Each 1-month increase in a child’s age increased the odds of receiving azithromycin treatment by 22% (aOR 1.22, 95% CI 1.15 - 1.30, *P*<0.0001). At the household-level, children whose head of household had a higher education (primary-level or above), experienced a higher-odds of receiving azithromycin compared to those reporting no education (aOR 1.77, 95% CI 1.04 - 3.02, *P* 0.04). Community population density was positively associated with receipt of treatment, with children living in more densely populated villages having a higher-odds of receiving azithromycin than children living in less densely populated communities (aOR 1.15, 95% CI 1.04 - 1.28, *P* 0.01). Child sex, community and household distance to the nearest primary health center, and number of children in the household were not found to be significantly related to child receipt of azithromycin in this survey. Results remained consistent in a sensitivity analysis that included Abalak CSI and children zero to one months of age (Supplementary Table 4).

**Table 2.**
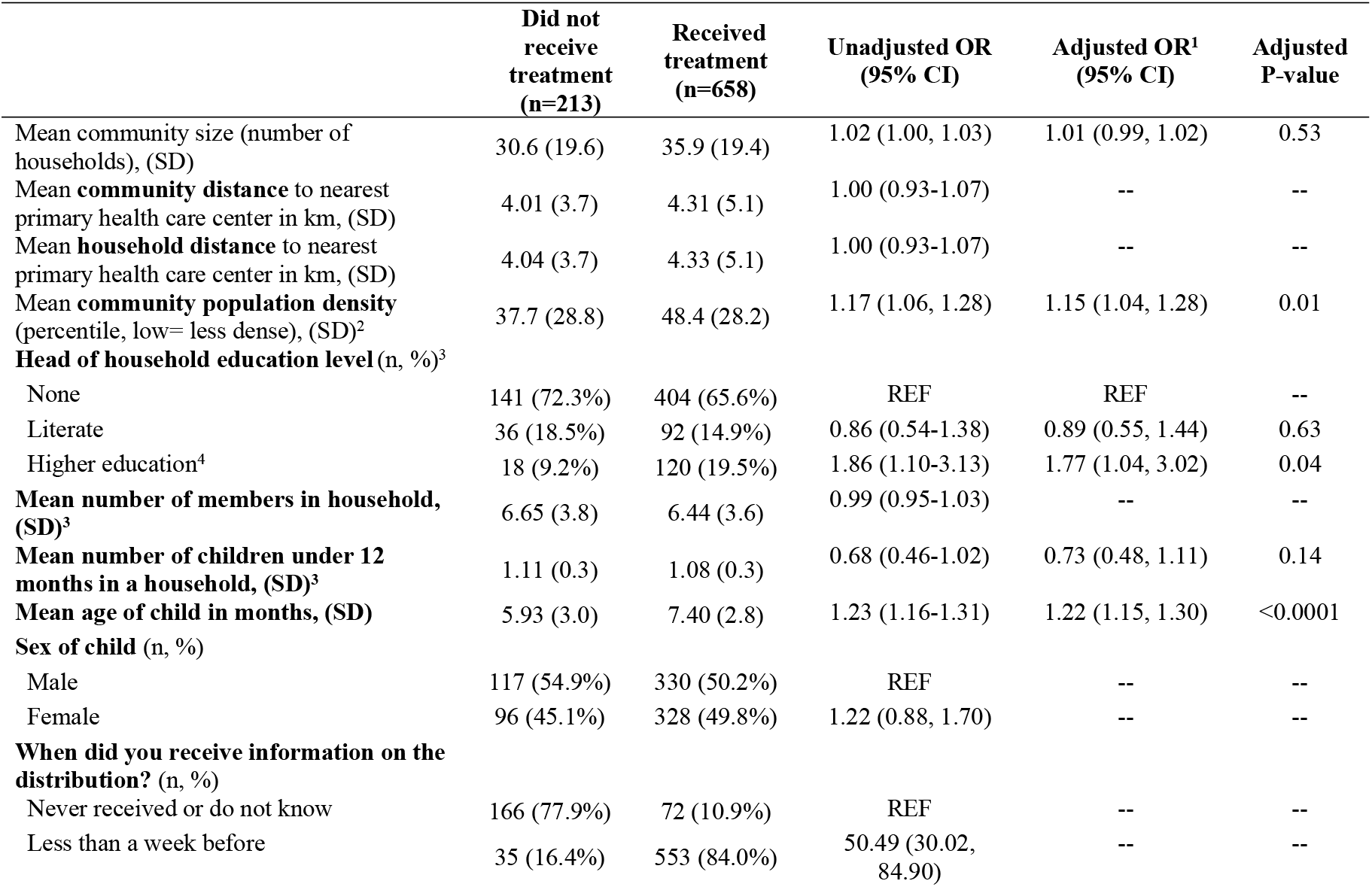
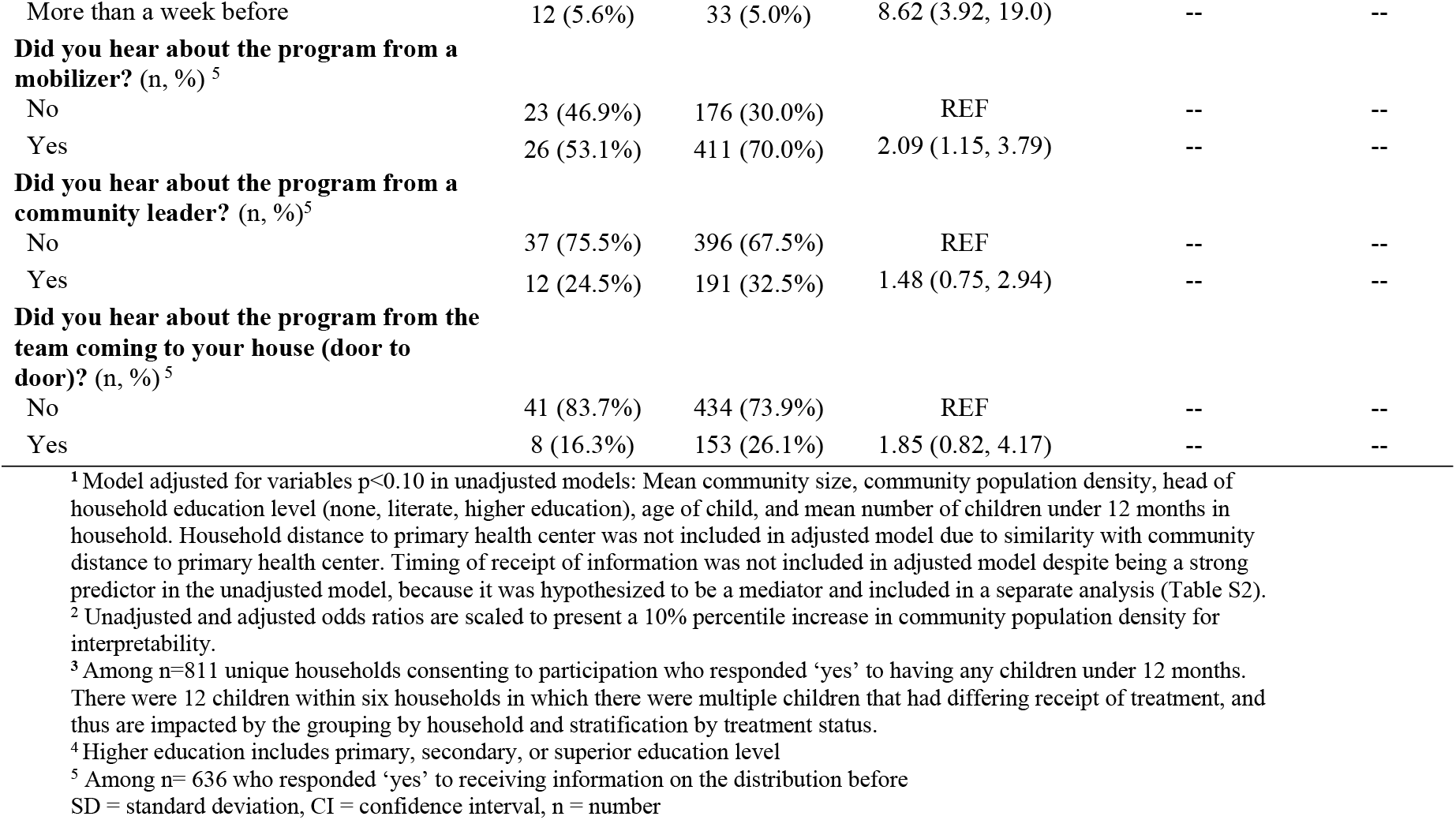
Unadjusted and adjusted odds ratios (OR) depicting the relationship between predictors and the receipt of treatment among households with at least one child under 12 months old (n=871). CSI Abalak and children 0-1 months of age are excluded.

**Fig 3.**
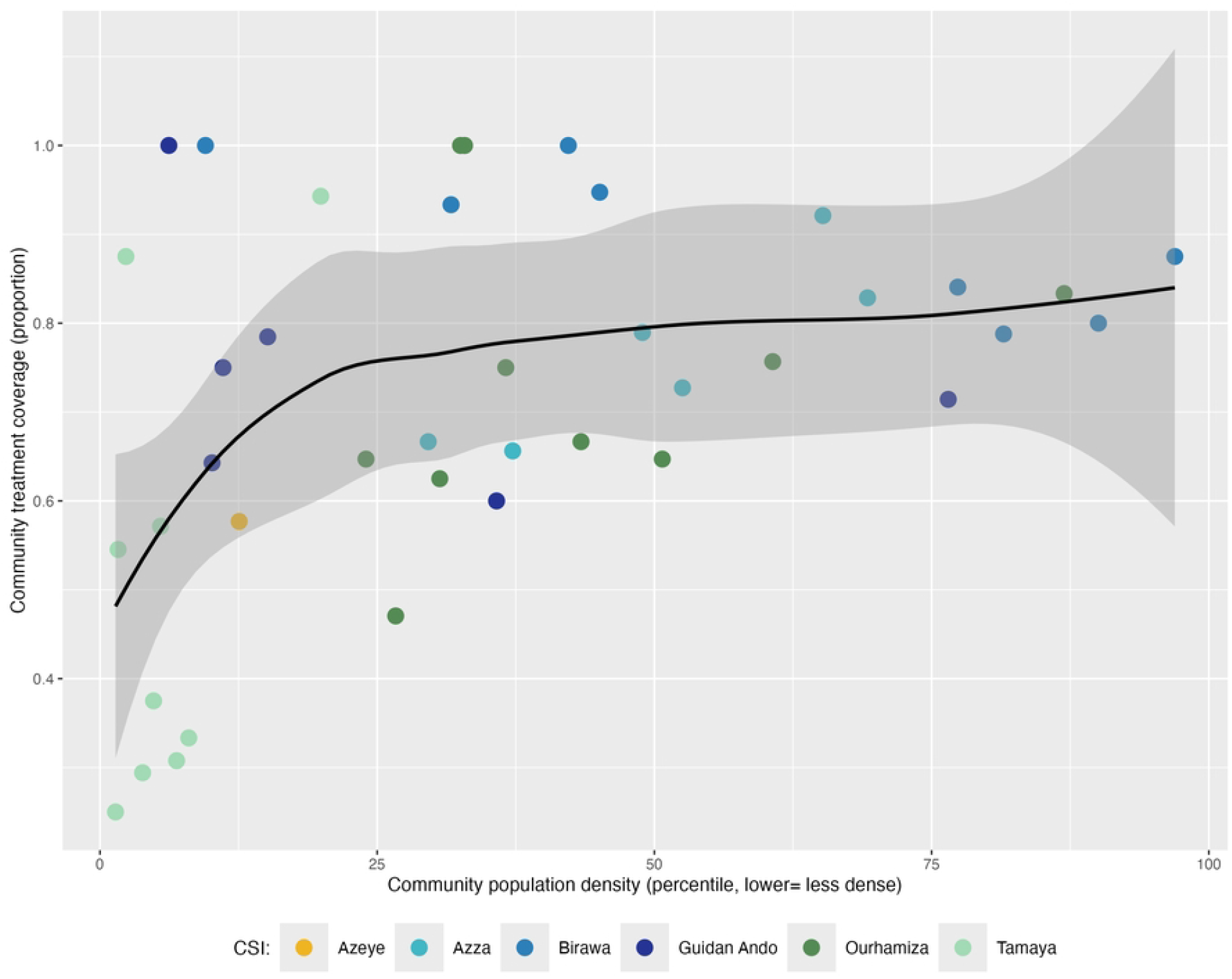
Community population density by community treatment coverage

Overall, 73.0% (636/871) of caregivers reported receiving information about the MDA in advance of the distribution (Table 1). Most caregivers 67.5% (588/871), reported receiving knowledge on the MDA distribution less than a week advance, and that group was 50.5 times more likely to participate in the MDA (95% CI 30.0 - 84.9) (Table 2). Age and population density were found to predict receipt of information about the MDA (Supplementary Table 3). Each 1-month increase in a child’s age increased the odds of receiving information about the treatment by 9% (aOR 1.09, 95% CI 1.03 - 1.15, *P* 0.003). Community population density was related to receipt of information, with more densely populated villages being more likely to have received information than communities which were less dense (aOR 1.16 - 95% CI 1.06, 1.27; *P* 0.001).

## Discussion

We conducted a coverage evaluation survey after azithromycin MDA for child survival to understand factors associated with non-participation in order to improve future implementation. Caregiver-reported coverage ranged from 60% to 86% across included CSIs, with the average CSI-level coverage of 72% being lower than both the 80% coverage target set for similar MDAs and the estimated administrative coverage of 96% using existing population denominators. Caregiver-reported coverage was lower in this study than in azithromycin MDA for child survival programs integrated with existing trachoma programs in Cote d’Ivoire and Nigeria,[9,10] although it was similar to estimates reported for azithromycin MDA for trachoma.[11] Indeed, we identified similar differences in caregiver-reported coverage and administrative coverage as comparisons reported for the trachoma program, which has found administrative sources to consistently overestimate coverage compared to self-reported sources.[11] Lymphatic filariasis and onchocerciasis MDA programs have also found similar differences between self-reported and administrative coverage estimates.[12,13] Although administrative estimates are highly feasible for programs to adopt, coverage evaluation surveys remain important additional sources of individual- and community-level information treatment uptake that can be used to improve implementation efforts.

The top three reasons reported by caregivers for their child not receiving azithromycin were absence during the distribution, nobody coming to the house, and not having received enough information about the distribution. Caregiver/child absence during the MDA was also among the most common reasons for non-participation in coverage surveys after azithromycin MDA for child survival in Nigeria and Cote d’Ivoire, suggesting that in these settings coordinated efforts to provide information on the timing of the MDA in advance and follow-up of missed households may be needed to increase coverage.[9] These findings align with previous surveys which demonstrated that drug-distributor no-shows[12,14] and lack of awareness[14] are barriers to participating in MDA. Notably, fear about side effects was not commonly reported as a reason for non-participation in this setting. Fear of side effects was consistently noted as a barrier to participation in some MDA programs like those for lymphatic filariasis,[15] but less so for azithromycin MDA done for trachoma or child survival.[9] Azithromycin is considered to be generally well tolerated with mild side effects, and adverse events have been uncommon in trials of azithromycin MDA for child survival.[3,16,17] In this programmatic setting, caregivers reported few adverse events, with less than 7% indicating any adverse event, and the most common being fever, diarrhea, and vomiting, consistent with the findings of prior studies.[9]

Advance receipt of information about the MDA was the strongest predictor of participation. Numerous other studies of azithromycin MDA and other MDA programs have identified advance knowledge of the campaign as an important predictor of participation.[18–21] In this setting, most caregivers reported receiving information on the MDA campaign less than a week in advance, which on its own was predictive of receiving treatment when compared to those who did not receive information before the distribution. Methods to transmit this information to communities included mobilizers, community leaders, and the team going door to door. Future sensitization campaigns would benefit from consistent advance notice of upcoming campaigns with reminders and targeting of groups likely to be missed.

Analyses identified younger age, less household education, and lower population density as other factors associated with non-participation in azithromycin MDA targeting children 1-11 months old, suggesting subgroups to target with concerted efforts to increase coverage. Younger children were less likely to participate than older children, which is largely consistent with findings from other MDA coverage surveys. [13,14,22] Caregivers may be hesitant to include the youngest children, underscoring the need for additional outreach and education efforts, especially since caregivers of younger children were also less likely to have received information about the distribution and the youngest age groups are at the highest risk of mortality.[3] Children residing in households where the head of household had no formal education were also less likely to have received treatment compared to those in households where the head of that household had at least a primary-level education. MDA coverage surveys in other settings have found that children in households where the adults had at least a primary or higher education (compared to no primary) were more likely to be treated.[7] Education level is closely linked to socioeconomic status, which has also been consistently associated with MDA participation, with higher socioeconomic status linked to higher participation.[12,23] Children living in less densely populated (more remote) communities were also less likely to receive azithromycin than those living in more densely populated areas. This program excluded urban settings, so these results are not comparable to those studies which have identified urban areas as more likely to experience low coverage.[22] Children in more remote or less densely populated areas may be missed because CHWs may not know where they live, especially in those cases where CHWs are not from the communities themselves. Additionally, reaching these populations takes additional time, and as MDA campaigns are often under pressure to complete the activity within a narrow time window, there may be less incentive to seek out these households. As the purpose of door-to-door distribution is to include all households, a key next step will be to develop strategies to ensure that children in less densely populated areas do not experience lower coverage.

Strengths of this study include the population-based design to ensure representativeness and incorporation of an existing survey tool that has been widely used to assess MDA coverage in other settings. However, this survey approach has several known limitations including recall bias and social desirability bias. We attempted to minimize the potential for recall bias by conducting the survey with in 4 weeks of treatment and showing the azithromycin bottle used to reduce confusion with other campaigns. Social desirability bias is plausible given the self-reported nature of the survey. Overall caregiver-reported coverage was lower than reported in other settings, somewhat mitigating this concern, though it is possible actual coverage was even lower. Conversely, caregivers may have underreported treatment receipt (stating they did not receive medicine when they actually did), if they were hoping that this response would result in an additional dose being provided. In addition, it is also possible that households that were absent during the MDA were also absent during the survey data collection, which would result in overestimation of participation. The survey included experienced census data collectors and maps to monitor completeness, so survey coverage of the population is likely to be higher than CHW coverage during the MDA. Finally, this study focused on characteristics of individuals, households, and communities and did not include assessment of program and delivery characteristics, which will be essential to understanding approaches to increasing coverage in Niger and across the region.

Overall, we found that caregiver-reported participation in azithromycin MDA for child survival was slightly lower than established targets for similar programs and lower than the administrative estimates. Standards for monitoring and evaluation of azithromycin MDA for child survival have not yet been developed, but coverage evaluation surveys should be considered for inclusion given the challenges in obtaining accurate coverage with administrative population estimates. Advance receipt of information on the MDA, age, education, and population density were key predictors of participation in the MDA, highlighting strategies to be developed to ensure high coverage and inclusion of the most vulnerable populations.

## Data Availability

Data can be accessed at publication at: https://osf.io/47jkd/overview?view_only=69ab394303594f00bf6c2eda662a5584

## Declarations

### Ethics approval and consent to participate

Approval for this trial was obtained from the Institutional Review Boards at the Niger Ministry of Health (*Comité Nationale Éthique pour la Recherche en Santé;* N 041/2020/CNERS) and the University of California, San Francisco (19-28387). Verbal consent was obtained from regional, district, health center, and community leaders for all study activities. At the household level, verbal consent was obtained from the head of household or the caregiver using an informed consent script prior to the initiation of the survey.

### Author contribution

AMA, RM, EL, TML, and KSO contributed to the conception and design of this study. AMA, AA, RM, AM, NG, ANH, DB, EL, BP, and CB contributed to the implementation of the study and acquisition of data. CB, AMA, RM, and KSO contributed to the analysis and interpretation of data. CB and KSO drafted the initial version of the manuscript and all authors critically reviewed, revised, and approved the final submitted manuscript. All authors have agreed to be accountable for their contributions and for the accuracy and integrity of the work.

## Acknowledgements

We would like to thank the many collaborators who have contributed to the AVENIR study group. **<u>Investigators and Study Personnel:</u> *Centre de recherche et interventions en santé publique, Niamey, Niger*** *–* Bawa Aichatou, Ahmed Arzika, Diallo Beidi, Ismael Mamane Bello, Fati Bello, Ousseini Boubacar, Nameywa Boubacar, Nasser Gallo, Karamatoulaye Hamadou, Amadou Harouna, Naser Harouna, Laminou Maliki Haroun, Alio Karamba, Mariama Keimago, Sani Mahamadou, Ramatou Maliki, Abarchi Moustapha, Abraham Omar, Farissatou Oumarou, Ismael Issa Sara**; *Programme national de santé oculaire, Niamey, Niger*** – Amza Abdou, Nassirou Beido, Boubacar Kadri, Boubacar Maïdanda**; *University of California, San Francisco, San Francisco, CA, USA*** – Benjamin Arnold, Cindi Chen, Emily Colby, Thuy Doan, Jeremy D Keenan, Sandrine Kyane, Elodie Lebas, Thomas M Lietman, Zijun Liu, William Nguyen, Kieran S O’Brien, Catherine E Oldenburg, Brittany Peterson, Travis C Porco, Kevin Ruder, George W Rutherford, Lina Zhong, Zhaoxia Zhou. **<u>Other Acknowledgements</u>: *Bill & Melinda Gates Foundation, Seattle, WA, USA*** – Rebecca Brander, Dennis Chao, James Heine, Rasa Izadnegahdar, Laura Lamberti, Assaf Oron, Surabhi Rajaram, Matthew Steele; ***Centre de recherche médical et sanitaire, Niamey, Niger*** *–* Ibrahima Issa, Ronan Jambou, Rabiou Labbo, S’Hooshim N. Lamine, Sani Ousmane, Boubakar Rakia, Maikano Sadikou; ***Centre de santé de la mère et de l’enfant, Dosso, Niger*** – Amina Seyfoulaye ; ***ClinEpiDB, Philadelphia, PA, USA*** *–*Brianna Lindsay, Nupur Kittur, David Roos, Sheena Tomko; ***Comité national éthique pour la recherche en santé, Niamey, Niger*** *–* Issa Adji, Djibo Ali, Souleymane Alzouma, Maidanda Boubacar, Diegou Boureima, Cheik Boureima Daouda, Idi Moussa Djatao, Ibrahim Jean Etienne, El Hadji Boubakar H Maiga, Amadou Oumarou, Ocquet Sakina, Sanoussi Samuila; ***Data and Safety Monitoring Committee*** – *Emory University, Task Force for Global Health –* David Addiss (chair); *Niger Ministry of Health* – Mourtala Assao; *St. George’s University of London* – Julia Bielicki, *Retired from CDC* – Allen Hightower, *Children’s Hospital of Colorado* – Brian Jackson, *University of Melbourne* – Fiona Russell**; *Global Health Strategies, New York, NY, USA*** – Frances Hocking, Zied Mhirsi, Dan Pawson**; *Pfizer, New York, NY, USA*** – Waqas Ahmed, Todd D. Hatajik, Marjan Javid, Julie Jenson, Chuck Knirsch, Chiao-Chin Lin; ***Speak Up Africa, Dakar, Senegal*** *–* Maelle Ba, Yaye Sophiétou Diop, Yacine Djibo, Gráinne Hutton, Fara Ndiaye**; *UCSF-UC Berkeley Center for Global Health Development, Diplomacy, and Economics, CA, USA*** *–* Elliot Marseille, Jim G. Khan, Stefano M. Bertozzi; ***University of Maryland School of Medicine*, Center for Vaccine Development and Global Health, *Baltimore, USA*** – Meagan Fitzpatrick.

